# Radiation associated brain image changes after proton therapy for skull base head and neck cancers

**DOI:** 10.1101/2020.02.06.20020610

**Authors:** Grete May Engeseth, Sonja Stieb, Abdallah Sherif Radwan Mohamed, Renjie He, Camilla Hanquist Stokkevåg, Marianne Brydøy, Clifton Dave Fuller, Adam S Garden, David I Rosenthal, Jack Phan, William H Morrison, Jay P Reddy, Richard Wu, Xiaodong Zhang, Steven Jay Frank, Gary Brandon Gunn

## Abstract

**Background and purpose:** To characterize patterns and outcomes of brain MR image changes after proton therapy (PT) for skull base head and neck cancer (HNC).

**Material and methods:** 127 patients treated with PT for HNC who had received at least 40 Gy(RBE) to the brain and had ≥ 1 follow-up MRI > 6 months after PT were analyzed. MRIs were reviewed for radiation-associated image changes (RAIC). MRIs were rigidly registered to planning CTs, and RAIC were contoured on T1 (post-contrast) and T2 weighted sequences, and dose-volume parameters extracted. Probability of RAIC over time was calculated using multistate analysis. Univariate/multivariate analyses were performed using Cox Regression. Recursive partitioning analysis was used to investigate dose-volume correlates of RAIC development.

**Results:** 17.3% developed RAIC. All RAIC events were asymptomatic and occurred in the temporal lobe (14), frontal lobe (6) and cerebellum (2). The median volume of the RAIC on post-contrast T1 was 0.5 cc at their maximum size. The RAIC spontaneously resolved in 27.3%, progressed in 27.3% and improved or were stable in 29.6% of patients. The 3-year actuarial rate of developing RAIC was 14.3%. Brain and RAIC lesion doses were generally higher for temporal lobe RAIC compared to frontal lobe RAIC. RAIC was observed in 63% of patients when V_67 Gy(RBE)_ of the brain ≥ 0.17 cc.

**Conclusion:** RAIC lesions after PT were asymptomatic and either resolved or regressed in the majority of the patients. The estimated dose–volume correlations confirm the importance of minimizing focal high doses to brain when achievable.

## Introduction

For head neck cancers (HNCs) at the skull base high doses can be delivered to the brain, potentially leading to radiation-associated image changes (RAIC) (1). Often the diagnosis is based solely on radiographic findings on post-treatment MRIs obtained for routine cancer surveillance purposes. For HNCs, information on RAIC stems largely from previous studies in patients with nasopharyngeal cancers (NPC), specifically temporal lobe RAIC. Following Intensity Modulated Radiation Therapy (IMRT), the reported incidence of temporal lobe RAIC ranges between 2.3% and 14.0%. The association between RAIC and brain dose is considered to be clear and D_max_, D1cc-D2cc and V_40 Gy(RBE)_ have been identified as important predictors. Analysis of dosimetric correlations with RAIC suggest that risk can be kept below 5% when doses of 63 Gy is delivered to small volumes of the brain (≤ 2cc), but increases towards 50% with doses of 70 Gy to 80 Gy (2–12).

Owing to the proximity to Central Nervous System (CNS) critical structures, HNC with tumor locations at the skull base or those with intracranial extension are often selected for treatment with proton therapy (PT) to improve or maintain target volume coverage while respecting normal tissues tolerance and reducing the toxicities and symptoms associated with the greater integral dose with IMRT. However, studies on the development of RAIC after PT for HNCs are limited, only a few mixed cohort studies exist where selected HNC are included, reporting crude RAIC rates of 18%-20% (13, 14). Thus, as part of a broader effort to better define the role and value of PT for HNC and to inform clinicians and the PT community regarding predictive toxicity modeling and to develop PT-specific dose constraints, the specific aims of this study were to:

1. Characterize the incidence and patterns of RAIC after PT for skull base HNCs
2. Identify candidate clinical and dose-volume parameters associated with RAIC
3. Propose practical dose constraints to minimize risk of RAIC

## Methods and materials

### Patient cohort

Adult patients treated with PT for HNC at University of Texas MD Anderson Cancer Center were eligible for participation in two consecutive Institutional Review Board-approved prospective studies (ClinicalTrials.org identifiers: NCT 00991094 and NCT 01627093). Participants with anatomic tumor location at the base of skull, including nasopharynx, sinonasal, periorbital, parotid, and skin and who had received a point dose of at least 40 Gy(RBE) to the brain and had ≥ 1 follow-up MRI > 6 months after completion of PT were considered eligible for this analysis. Those with prior radiation therapy to the region or who had received photon-based treatment as part of therapy were excluded.

### Treatment

All PT plans were based on non-contrast CT images and generated in the Eclipse treatment planning system (Varian Medical Systems, Palo Alto, CA, USA) using either Intensity Modulated Proton Therapy (IMPT), Passive Scattered Proton Therapy (PSPT) or a combination of IMPT/PSPT. For dose prescription a proton relative biological effectiveness (RBE) value of 1.1 was used as recommended by the ICRU (15). Dose (prescribed to CTV) and fractionation regimens were generally 60 Gy(RBE) in 30 fractions or 63-66 Gy(RBE) in 30-33 fractions in the postoperative PT setting or 66-70 Gy(RBE) in 33-35 fractions in the definitive PT setting. Induction chemotherapy regimens used were typically 3 cycles with taxane and platinum or taxane, platinum and fluorouracil (TPF), and concurrent chemotherapy was usually platinum-based.

### Follow-up and diagnosis of RAIC

Initial post-PT evaluations took place at 8 to 12 weeks after therapy completion and then every 3 months in the first year, every 4 months in the second year, every 6 months until 5 years, and annually thereafter. Diagnosis of RAIC was based on the following criteria: newly developed image changes in terms of i) contrast enhanced lesions on T1 weighted MR images and/or ii) cysts or white matter lesions manifested as high signal intensity on the T2 weighted sequences (16). All post-treatment diagnostic MRI reports and all images were individually reviewed for RAIC. No additional RAIC to those already diagnosed in the MRI reports was discovered. The clinical grading of RAIC was according to CTCAE v4.03 (17) (Supplementary Table I).

### Extraction of clinical and treatment related data

Patient, tumor and treatment characteristics were retrieved from the patients’ medical records (Epic Systems Corporation, Verona, WI) and the treatment planning system. The MRIs were rigid registered to the initial treatment planning CT and qualitatively evaluated through visual inspection and further manually corrected if deemed necessary. Gadolinium contrast enhanced lesions on T1 sequences (T1 lesions) and hyperintensities on T2 sequences (T2 lesions) were contoured as individual structures and propagated to the planning CT. Dose-volume histograms (DVHs) for all patients were exported using bin size of 0.1 Gy(RBE) from where relevant dose volume parameters for the brain tissue were retrieved, including: the maximum dose (D_max_), the dose delivered to 0.5cc (D_0.5cc_) and 1cc (D_1cc_) to 5cc (D_5cc_) of the brain in 1cc steps, as well as the volume of brain receiving 40-70 Gy(RBE) in 1 Gy(RBE) steps (V_40Gy(RBE_) to (V_70Gy(RBE)_). In addition, for each patient the T1 and T2 lesion volumes with the corresponding minimum dose (D_min_), the mean dose (D_mean_) and the D_max_ were extracted for the first MRI and the MRI with the largest volume of RAIC (“worst” MRI).

### Statistical analysis

The follow-up time was calculated as the time between the last PT fraction and the patient’s last follow-up MRI, the RAIC latency time as time between last PT fraction and first MRI with RAIC. Time to resolution from RAIC was calculated from date of the first MRI with RAIC and the date of the first MRI where lesion had resolved. Patients without RAIC at last follow-up MRI were censored at this time point.

Typical evolution for RAIC is an initial progressing phase, a regression phase and in some cases complete resolution (18). To account for recovered RAIC we applied a multistate survival analysis to calculate the probabilities of having RAIC at different time points (19, 20).

Comparisons between patients with and without RAIC were performed using the Mann Whitney U test, the Chi Square and Fishers exact test. All tests were two-sided with a significance level of 0.05. Correlations between the clinical and dosimetric predictor variables were calculated using Spearman’s rank correlation and Pearson’s correlation coefficient. Univariate and multivariate analysis of the association between RAIC and candidate predictors were performed using Cox Proportional Hazards Regression Analysis (21, 22). Covariates with p value < 0.2 from the univariate analysis were considered for inclusion in the multivariate analysis. However, since DVH parameters are highly correlated, the dose volume variables with the lowest significance level from variables where the correlation coefficient were > 0.8 were chosen for the analysis.

To identify practical dose constraints, recursive partitioning analysis (RPA) was applied. RAIC occurring within three years after treatment was chosen as endpoint; therefore dose volume statistics for patients who developed RAIC within three years post PT as well as patients without RAIC who had a minimum of 36 months of follow-up formed the data material. The tree growing criteria were as follows: number of observations in a terminal node was constrained to 10% of the dataset and the splitting was based on the Gini index. Bootstrapping (number of resampling = 1000) was used for estimating model uncertainty (23, 24). Statistical analyses were performed in SPSS version 24 (IBM SPSS Statistics for Windows, Version 24.0. Armonk, NY: IBM Corp, US) and R version 3.6 (R Foundation for Statistical Computing, Vienna, Austria).

## Results

### Patient and treatment characteristics

Three hundred and seven patients treated from 12/2010 through 06/2018 with the eligible anatomic tumor location were reviewed. Of these, 66 were excluded due to prior photon treatment to the same region, 78 due to no post-treatment MRI 6 > months, and 36 due to brain dose < 40 Gy(RBE). The remaining 127 patients formed the cohort.

Patient, tumor and treatment characteristics are displayed in Table I. 85 patients (66.9%) were treated exclusively with IMPT, 15 patients (11.8%) exclusively with PSPT and 27 patients (21.3%) with a combination of PSPT and IMPT. Thirty eight patients (29.9%) had one adaption of the treatment plan and six patients (4.7%) had two. Twenty patients (15.7%) received both induction and concurrent chemotherapy.

**Table I:** Patient, treatment and tumor characteristics for the entire cohort and by RAIC group

| Characteristic | All patients (n = 127) |  | No RAIC (n = 105) |  | RAIC (n = 22) |  | p |
| --- | --- | --- | --- | --- | --- | --- | --- |
|  | n | % | n | % | n | % |  |
| **Sex** |  |  |  |  |  |  | **0.98** |
| Male | 67 | 52.8 | 57 | 54.3 | 10 | 45.5 |  |
| Female | 60 | 47.2 | 48 | 45.7 | 12 | 54.3 |  |
| **Age** |  |  |  |  |  |  | **0.69** |
| Median (Q1-Q3) | 51 (39-64) |  | 50 (37-63) |  | 54 (41-61) |  |  |
| **Anatomic Site** |  |  |  |  |  |  | **0.16** |
| Base of Skull, NOS | 7 | 6.3 | 6 | 5.7 | 1 | 4.5 |  |
| Nasopharynx | 35 | 26.8 | 27 | 25.7 | 8 | 36.4 |  |
| Sinonasal | 29 | 22.8 | 20 | 19.0 | 9 | 40.9 |  |
| Parotid | 14 | 11 | 14 | 13.3 | - | - |  |
| Orbital | 23 | 18.1 | 21 | 20.0 | 2 | 9.1 |  |
| Skin | 10 | 7.9 | 9 | 8.6 | 1 | 4.5 |  |
| Other | 9 | 7.1 | 8 | 7.6 | 1 | 4.5 |  |
| **T-category** |  |  |  |  |  |  | **0.77** |
| T1-T2 | 28 | 22 | 24 | 22.9 | 4 | 18.2 |  |
| T3-T4 | 53 | 41.7 | 41 | 39 | 12 | 54.5 |  |
| Recurrent | 29 | 22.8 | 25 | 23.8 | 4 | 18.2 |  |
| Tx | 5 | 3.9 | 5 | 4.8 | - | - |  |
| No stage | 12 | 9.4 | 10 | 9.5 | 2 | 9.1 |  |
| **Surgery** |  |  |  |  |  |  | **0.23** |
| No | 49 | 38.6 | 38 | 36.2 | 11 | 50 |  |
| Yes | 78 | 61.4 | 67 | 63.8 | 11 | 50 |  |
| **Induction chemotherapy** |  |  |  |  |  |  | **0.76** |
| No | 106 | 83.5 | 86 | 81.9 | 20 | 90.9 |  |
| Yes | 21 | 16.5 | 19 | 18.1 | 2 | 9.1 |  |
| **Concurrent chemotherapy** |  |  |  |  |  |  | **0.04** |
| No | 54 | 42.5 | 49 | 46.7 | 5 | 22.7 |  |
| Yes | 73 | 57.5 | 56 | 53.3 | 17 | 77.3 |  |
| **PT technique** |  |  |  |  |  |  | **0.72** |
| IMPT | 85 | 69.9 | 69 | 65.7 | 16 | 72.7 |  |
| PSPT | 15 | 11.8 | 12 | 11.4 | 3 | 13.6 |  |
| IMPT/PSPT | 27 | 21.3 | 24 | 22.9 | 3 | 13.6 |  |
| **Prescribed dose** |  |  |  |  |  |  |  |
| Median (Q1-Q3) | 64.0 (60.1-69.7) |  | 64.0 (60.1-68.3) |  | 69.3 (63.1-70.0) |  | **0.04** |
| **Brain D<sub>max</sub> < 60 Gy(RBE)** |  |  |  |  |  |  | **0.01** |
| No | 95 | 74.8 | 74 | 70.5 | 21 | 96.9 |  |
| Yes | 32 | 25.2 | 31 | 29.5 | 1 | 3.1 |  |
| **Brain D<sub>max</sub> > 70 Gy(RBE)** |  |  |  |  |  |  | **< 0.01** |
| No | 89 | 70.1 | 81 | 77.1 | 8 | 36.4 |  |
| Yes | 38 | 29.9 | 24 | 22.9 | 14 | 63.6 |  |

Q1: 25 percentile, Q3: 75 percentile. Base of Skull: Adenoid cystic carcinoma, adenocarcinoma, neuro-endocrine carcinoma, chondrosarcoma, ameloblastoma, osteosarcoma. Other: vagus nerve schwannoma, paraganglioma, melanoma inferior turbinate, unknown primary site. IMPT: Intensity Modulated Proton Therapy, PSPT: Passive Scattering Proton Therapy

### Outcome and patterns of RAIC

Median follow-up time was 29 months (range: 6-97). Twenty-two patients (17.3%) developed RAIC with a median latency of 24 months (9-37); all but one RAIC had developed within three years after PT. Median follow-up time after diagnosis of RAIC was 14 months (0-70). All cases of RAIC had been incidentally discovered on routine surveillance imaging and all patients were asymptomatic at initial diagnoses and subsequent follow-up visits. However, due to progression of the image changes on the subsequent MRIs two patients were treated with pentoxifylline and vitamin E, one of whom also received dexamethasone, and one patient was treated with bevacizumab (25). Therefore, 19 patients were considered to have Grade 1 and three patients Grade 3 CNS necrosis according to CTCAE 4.03.

At last follow-up MRI the lesions had resolved in six patients, progressed in four, regressed in four, were stable in four and four patients had yet to have a follow-up MRI. A case example of the observed RAIC evolution including overview of evolution timeline can be found in Supplementary Material Figure 1–2. Overall, the estimated probability of RAIC at two, three and five years was 8.7 % (95% CI: 3.8%, 13.7%), 14.3% (95% CI: 8.2%, 20.4%), and 12.7% (95% CI: 6.9%, 18.5%), respectively (Figure 1).

**Figure 1:**
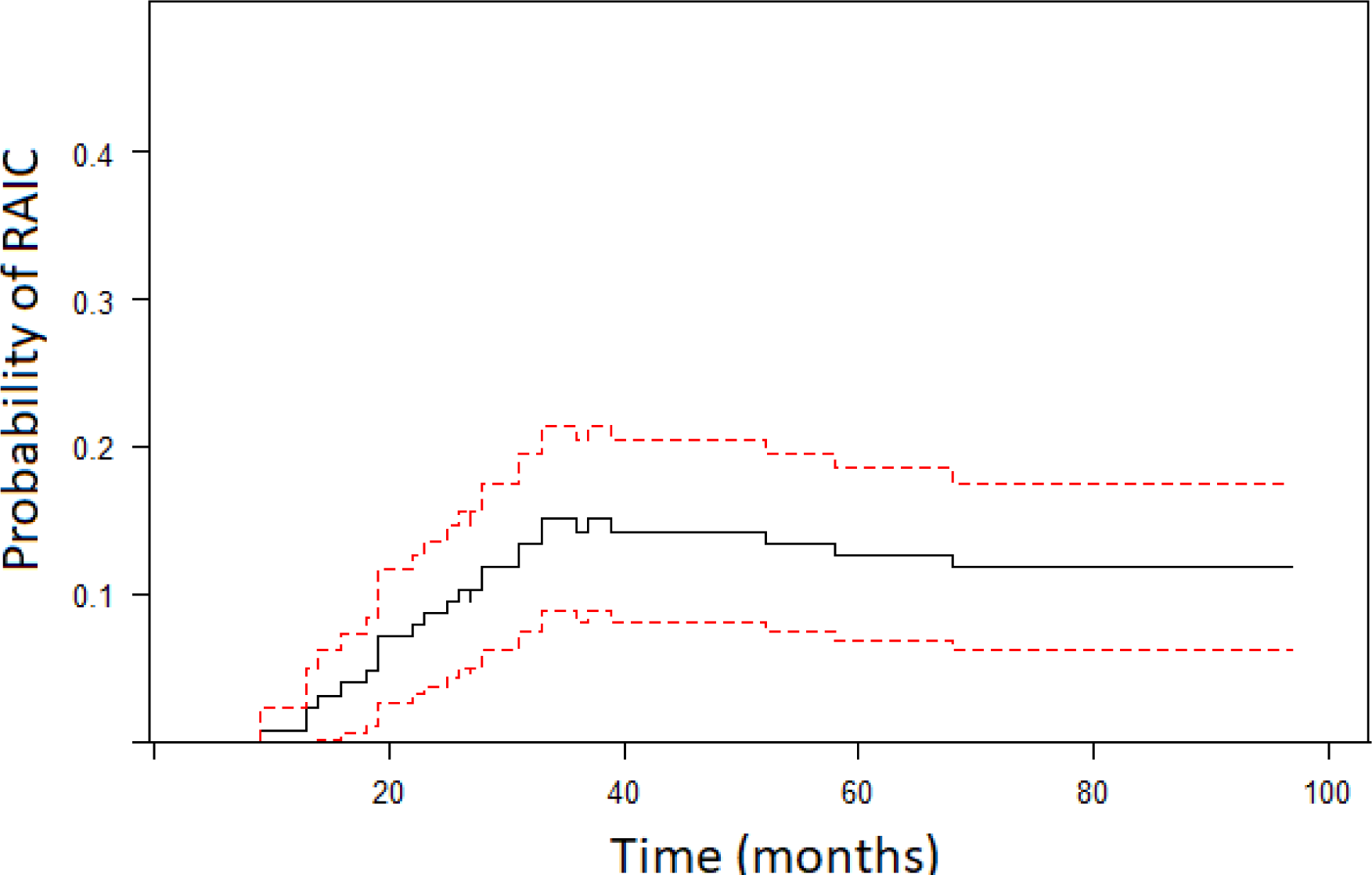
The probability of having RAIC over time (black solid line) including 95% confidence intervals (red dotted lines).

**Figure 2:**
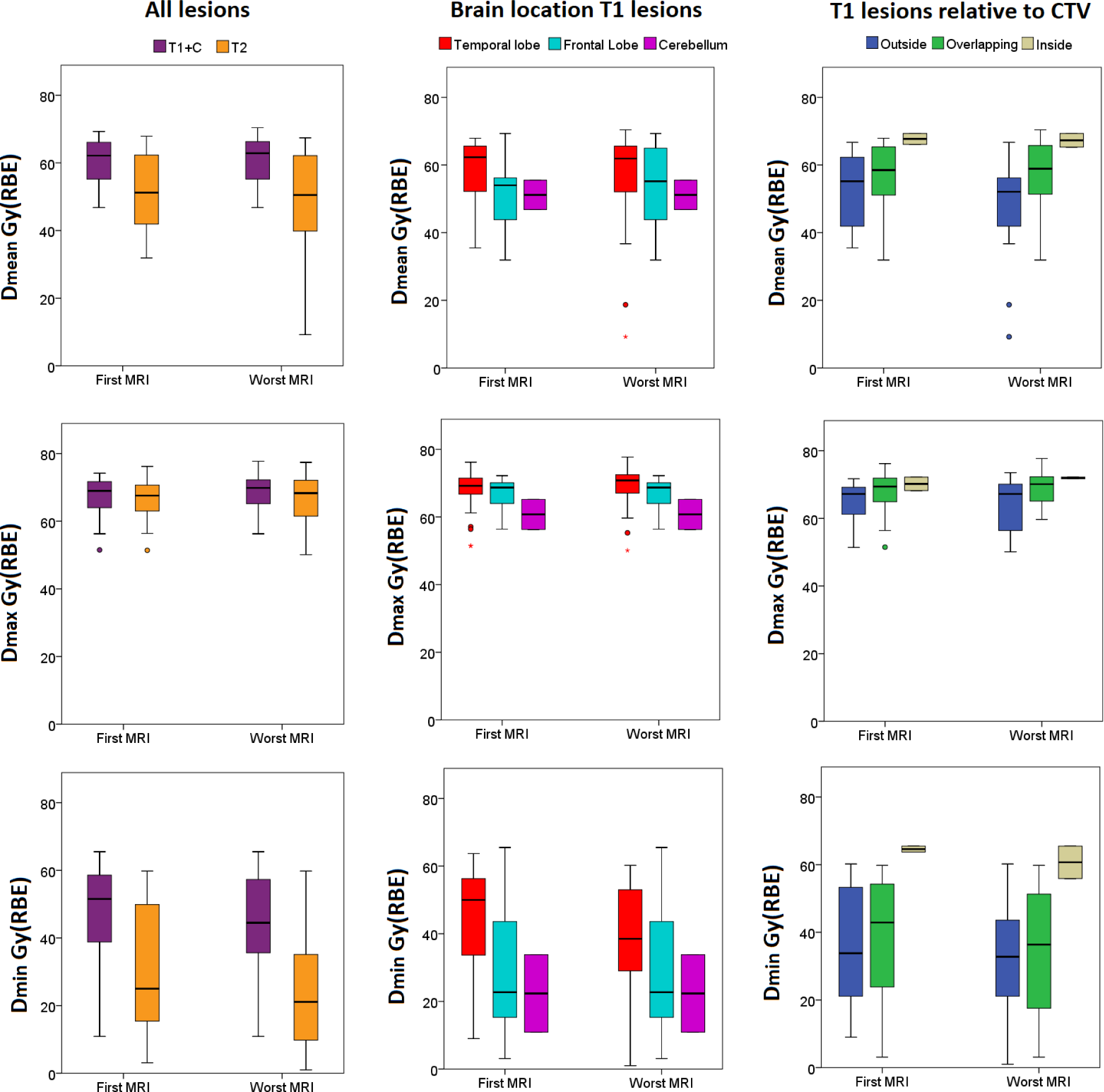
RAIC lesion doses for all lesions (left column), according to location in the brain (middle column) and location relative to the CTV.

Tumor and treatment characteristics for patients with RAIC including dose-volume summary statistics for the contoured lesions are provided in Supplementary Materials (Table II-VI). 68.2% patients diagnosed with RAIC had been treated for T4 or unresectable disease, and 63.6% had intracranial tumor extension. RAIC was located in the temporal lobe (14 patients), frontal lobe (six patients) and in the cerebellum (two patients). Prescribed CTV dose and brain doses were numerically higher in patients with RAIC in the temporal lobe compared to those with RAIC in the frontal lobe and cerebellum (Table II).

**Table II:** Prescribed CTV dose and brain doses for patients with RAIC are displayed according to RAIC location in the brain. Median values and Q1 to Q3 is reported. Reported dose and volumes in Gy(RBE) and cc, respectively.

| CTV/ brain dose- volume parameters | RAIC location |  |  |
| --- | --- | --- | --- |
|  | Temporal lobe | Frontal lobe | Cerebellum |
| Prescribed CTV dose | 70.0 (68.5-70.1) | 60.0 (60.0-68.2) | 60.0 (-) |
| D<sub>max</sub> | 74.7 (69.5-77.7) | 70.3 (64.6-73.8) | 64.5 (-) |
| D<sub>1cc</sub> | 71.4 (64.5-73.0) | 63.9 (61.3-68.3) | 61.7 (-) |
| D<sub>2cc</sub> | 68.9 (61.0-72.2) | 62.7 (58.4-67.7) | 60.8 (-) |
| V<sub>40 Gy(RBE)</sub> | 48.9 (33.0-71.5) | 26.9 (17.6-41.4) | 43.6 (-) |
| V<sub>50 Gy(RBE)</sub> | 25.1 (15.9-37.0) | 15.2 (9.4-24.9) | 27.4 (-) |
| V<sub>60 Gy(RBE)</sub> | 10.0 (2.5-17.9) | 6.1 (1.2-10.8) | 9.3 (-) |
| V<sub>70 Gy(RBE)</sub> | 1.6 (0.0-5.5) | 0.0 (0.0-0.2) | 0.0 (-) |

The majority of RAIC developed in patients with nasopharyngeal or sinonasal cancers (77.3%); for the patients with sinonasal cancer RAIC was found both in the temporal lobe (four) and the frontal lobe (five), whereas all (eight) RAIC occurred in the temporal lobe for the nasopharyngeal cancers (Supplementary Material Figure 3). Two of the contrast-enhanced T1 lesions were inside and seven were outside the CTV, the remaining 13 were overlapping with the CTV (Supplementary Material Figure 4). At first MRI 92% of the T1 contrast-enhanced lesions were 1cc or less, with a median volume of 0.3cc (punctuate to 1.1cc); on the MRIs where the RAIC was at its largest size, 77% of the T1 lesions were less than 1cc with a median volume of 0.5cc (0.1cc to 3.1cc) (Supplementary Material Table III-V). The T1 lesion doses were highest in lesions located in the temporal lobe and for lesions inside the CTV (Figure 2).

**Figure 3:**
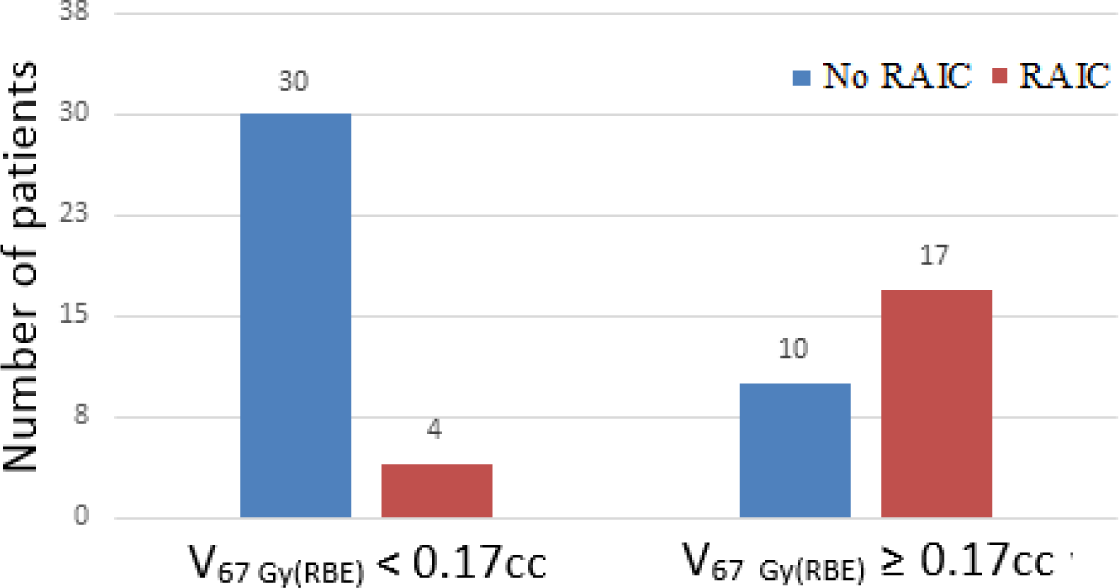
Results from RPA: 17 out of 27 patients where V_67 Gy(RBE)_ ≥ 0.17cc and four out of 30 patients with V_67 Gy(RBE)_ < 0.17cc developed RAIC, respectively.

### Clinical and dose volume correlations with RAIC

Statistical significant differences by RAIC group were only found for prescribed dose and brain dose-volume parameters (Table I/ Supplementary Material Table VII). V_40 Gy(RBE)_-V_70 Gy(RBE)_ for the brain were included in the RPA. The final model was a one leaf tree with splitting of V_67 Gy(RBE)_ at 0.17cc; 81% of the patients with RAIC had a V_67 Gy(RBE)_ ≥ 0.17cc and 63.1 % of the patients with V_67 Gy(RBE)_ ≥ 0.17cc developed RAIC (Figure 3 /Supplementary Material Table VIII). Based on the result from RPA, we included V_67 Gy(RBE)_ ≥ 0.17cc in the univariate cox regression analysis. From this analysis prescribed dose, D_max_, D_1-5cc_, V_70 (GyRBE)_ and V_67 Gy(RBE)_ ≥ 0.17cc were significantly associated with the development of RAIC. In the multivariate analysis; V_67 Gy(RBE)_ ≥ 0.17cc was the only statistically significant predictive factor (HR: 7.57, 95% CI: 1.48, 38.76, p = 0.015) (Table III).

**Table III:** Results from univariate and multivariate cox regression analysis of clinical and dosimetric variables associated with development of RAIC.

|  | Univariate analysis |  | Multivariate analysis |  |
| --- | --- | --- | --- | --- |
|  | HR (95% CI) | *p* | HR (95% CI) | *p* |
| Age | 1.02 (0.99-1.04) | > 0.1 |  |  |
| Anatomical site* | 2.52 (0.99-6.45) | 0.054 | 1.23 (0.41-4.05) | > 0.1 |
| T3-T4** | 1.83 (0.79-4.25) | > 0.1 | 0.44 (0.12-1.66) | > 0.1 |
| Surgery | 0.56 (0.24-1.30) | > 0.1 | 2.46 (0.41-4.05) | > 0.1 |
| Induction chemotherapy | 0.67 (0.20-2.28) | > 0.1 |  |  |
| Concurrent chemotherapy | 2.22 (0.82-6.03) | > 0.1 | 1.99 (0.42-9.33) | > 0.1 |
| PT technique*** | 1.06 (0.41-2.72) | > 0.1 |  |  |
| Prescribed dose | 1.11 (1.01-1.23) | 0.035 | 1.03 (0.88-1.20) | > 0.1 |
| D<sub>max</sub> | 1.15 (1.07-1.24) | < 0.01 | 1.06 (0.92-1.21) | > 0.1 |
| D1<sub>cc</sub> | 1.11 (1.05-1.18) | < 0.01 |  |  |
| D2<sub>cc</sub> | 1.10 (1.04-1.16) | < 0.01 |  |  |
| D3<sub>cc</sub> | 1.08 (1.03-1.13) | < 0.01 |  |  |
| D4<sub>cc</sub> | 1.07 (1.02-1.12) | < 0.01 |  |  |
| D5<sub>cc</sub> | 1.07 (1.02-1.12) | < 0.01 |  |  |
| V<sub>40</sub> GY(RBE) | 1.01 (0.998-1.02) | 0.061 | 0.99 (0.98-1.01) | > 0.1 |
| V<sub>50</sub> GY(RBE) | 1.01 (0.996-1.03) | 0.080 |  |  |
| V<sub>60</sub> GY(RBE) | 1.01 (0.985-1.04) | > 0.1 |  |  |
| V<sub>70</sub> GY(RBE) | 1.18 (1.03-1.34) | < 0.01 | 1.06 (0.87-1.29) | > 0.1 |
| V<sub>67</sub> GY(RBE) ≥ 0.17<sub>cc</sub> | 7.72 (2.84-21.0) | < 0.01 | 7.41 (1.48-38.76) | 0.015 |

\* Nasopharyngeal carcinoma/sinonasal vs other, \*\*T3/T4 vs other. \*\*\* IMPT vs other, HR: Hazard Ratio

## Discussion

In the current study, we characterized the patterns and clinical outcomes of RAIC after PT in a relatively large and heterogeneous cohort of patients with HNCs at the skull base. The crude incidence rate of RAIC in our study was 17.3%, but given the transient nature of observed lesions, the estimated probability of RAIC at 2, 3, and 5-years was 9%, 14%, and 13%, respectively.

All patients with RAIC in our study were asymptomatic. Our findings are in contrast to the higher proportions of patients with symptomatic RAIC reported in other studies (Supplementary Materials Table IX), where reported rates of clinical symptoms amongst patients with RAIC range between 22% and 47% for IMRT cohorts (2, 3, 5, 10, 12), and 17% and 42% for proton cohorts (13, 14, 26). There are several possible explanations for our findings in contradistinction to other series. Both higher doses and larger RAIC volumes have been suggested to have an impact on the development of clinical symptoms (1, 25, 27). In the current study the median T1 and T2 lesion volumes at its worst were 0.5cc and 2.9cc, respectively, comparatively smaller than reported elsewhere. Zhou et al (12) reported 30% clinical symptoms in a large series (1887) of NPC patients receiving IMRT and considerable larger mean T1 and T2 lesion volumes of 4.2cc and 21.7cc, respectively. Su et al (6) reported mean T1 and T2 lesion volumes of 7.7cc and 21.2cc and 22% clinical symptoms. None of these studies, however, analyzed the correlation between lesion volume and clinical symptoms. McDonald et al. (13) found no significant difference in RAIC lesion volumes between asymptomatic and symptomatic patients (Grade 1 vs. Grade 2) in their series of skull base cancers treated with PSPT, however a significant difference in the mean and maximum lesion doses was found. A second explanation is that asymptomatic RAIC could be considered an early sign of impending symptomatic progression, future development of clinical symptoms for those with stable, no follow-up MRI or progressing RAIC in our cohort cannot be ruled out.

Estimated dose-volume constraints from previous studies on HNC IMRT cohorts are fairly consistent emphasizing the importance of avoiding a focal high dose to the brain, although a few studies have also showed increased probability of RAIC when V_40-45_ Gy of the temporal lobe increases (Supplementary Table IX). In our analysis V_67 Gy(RBE)_ ≥ 0.17 cc was identified as a predictor of RAIC development, both based the Cox Regression analysis (HR 7.41) and on RPA. From RPA we found that V_67 Gy(RBE)_ less than 0.17cc resulted in 12% of patients developing RAIC and 63% when V_67 Gy(RBE)_ ≥ 0.17cc. Huang et al (3) reported a 13% and 63% probability of temporal lobe RAIC with D1cc < 71.1 Gy and D1cc > 71.1 Gy respectively, in large cohort of nasopharyngeal cancers treated with IMRT. From the dose-response analysis of 66 skull base patients treated with PSPT, McDonald et al. (13) reported a probability of 15% temporal lobe RAIC when V_70 Gy(RBE)_ < 1.7cc.

PT-specific uncertainties caused by range uncertainty, elevated linear energy transfer (LET) and the variable RBE of protons, and their effect on the dose distribution and RAIC development has not been addressed in this study. The majority of the RAIC lesions were outside or overlapping with the CTVs and thus located in the area where the effect of proton range uncertainty or the increase in RBE/LET is expected to be largest. The potential LET/RBE correlations with the development of RAIC has indeed been demonstrated for intracranial tumors in pediatric ependymoma patients (28), pediatric brain tumors (29) and recently in a small patient cohort of glioma tumors where, similar to the current study, the lesions were located at border of the CTV (30).

Since V_40 Gy_ to the brain has previously been identified as predictor of RAIC, our brain dose threshold for including patients was set to 40 Gy(RBE). Our study adds to existing literature in that we included multiple HNC disease sites and investigated RAIC in any lobe of the brain. The radiosensitivity of the different lobes may vary, as will the symptom constellation and clinical significance of symptomatic RAIC. Our dosimetric analysis showed higher brain- and RAIC lesion doses for lesions located in the temporal lobes compared to those in the frontal lobe, which may indicate that the temporal lobe is less radiosensitive than the frontal lobe in terms of developing RAIC, an observation worthy of additional investigation.

Although all MRI reports and images in the current study were retrospectively reviewed and diagnoses verified, inherent limitations of retrospective review apply. Other limitations in our study exist, including a relatively short follow-up time in our cohort, as RAIC can develop at a later time point or existing lesions could potentially progress. The MRIs in our study were acquired for standard post treatment surveillance purposes, and T2 flair sequences were not available, thus standard T2 sequences were used for contouring T2 edema signal, which can be less accurate than T2 flair. Further, since the lesions were very small, accuracy of the calculation of the lesion doses may be influenced by uncertainties in the image registration procedure. Finally, our material included a variety of tumor sites treated with both PSPT and IMPT. PSPT generally has a larger irradiated volume, whereas IMPT is conformal, but more sensitive to uncertainties. Although PT technique was not significantly correlated with RAIC development, we cannot rule out that it may influence our results. In addition, the study could also have limited power to define dose-volume constraints since our cohort was too small to split into a training and a validation set for RPA. Therefore, our results should be externally validated.

## Conclusion

RAIC lesions after PT for skull base HNC were small, asymptomatic and regressed or completely resolved in the majority of patients affected. Our dosimetric analysis suggest that the temporal lobes could be more radio-resistant than the frontal lobe. The estimated dose–volume correlations identified in our study confirm the importance of minimizing focal high doses to brain (V_67 Gy(RBE)_ < 0.17 cc) when achievable. We are presently exploring the potential impact of LET distribution and variable RBE on RAIC development.

## Data Availability

The authors confirm that the data supporting the findings of this study are available within the article and its supplementary materials.

## Notes

### Competing Interest Statement

All authors have completed the ICMJE uniform disclosure form and declare: Engeseth is funded by Trond Mohn Foundation. Dr S Stieb is funded by the Swiss Cancer League (BIL KLS-4300-08-2017). Stokkevåg is funded by Kreftforeningen. Dr Fuller received funding and salary support related to this project from the National Institutes of Health (NIH), including: the National Institute for Dental and Craniofacial Research Establishing Outcome Measures Award (1R01DE025248/R56DE025248) and an Academic Industrial Partnership Grant (R01DE028290). Dr. Fuller received funding and salary support unrelated to this during the project from: National Science Foundation (NSF), Division of Mathematical Sciences, Joint NIH/NSF Initiative on Quantitative Approaches to Biomedical Big Data (QuBBD) Grant (NSF 1557679); a National Institute of Biomedical Imaging and Bioengineering (NIBIB) Research Education Programs for Residents and Clinical Fellows Grant (R25EB025787-01); the NIH Big Data to Knowledge (BD2K) Program of the National Cancer Institute (NCI) Early Stage Development of Technologies in Biomedical Computing, Informatics, and Big Data Science Award (1R01CA214825); NCI Early Phase Clinical Trials in Imaging and Image-Guided Interventions Program (1R01CA218148); an NIH/NCI Cancer Center Support Grant (CCSG) Pilot Research Program Award from the UT MD Anderson CCSG Radiation Oncology and Cancer Imaging Program (P30CA016672) and an NIH/NCI Head and Neck Specialized Programs of Research Excellence (SPORE) Developmental Research Program Award (P50 CA097007). Dr. Fuller has received direct industry grant support, honoraria, and travel funding from Elekta AB. S.J. Dr. Frank reports personal fees from Varian, grants and personal fees from C4 Imaging, grants from Eli Lilly, grants from Elekta, grants and personal fees from Hitachi, other from Breakthrough Chronic Care, personal fees from Boston Scientific, and personal fees from National Comprehensive Cancer Center (NCCN). Dr. Gunn reports philanthropic donation from the Family of Paul W. Beach.

### Clinical Trial

NCT 00991094, NCT 01627093

### Funding Statement

Engeseth is funded by Trond Mohn Foundation. Dr S Stieb is funded by the Swiss Cancer League (BIL KLS-4300-08-2017). Stokkevåg is funded by Kreftforeningen. Dr Fuller received funding and salary support related to this project from the National Institutes of Health (NIH), including: the National Institute for Dental and Craniofacial Research Establishing Outcome Measures Award (1R01DE025248/R56DE025248) and an Academic Industrial Partnership Grant (R01DE028290). Dr. Fuller received funding and salary support unrelated to this during the project from: National Science Foundation (NSF), Division of Mathematical Sciences, Joint NIH/NSF Initiative on Quantitative Approaches to Biomedical Big Data (QuBBD) Grant (NSF 1557679); a National Institute of Biomedical Imaging and Bioengineering (NIBIB) Research Education Programs for Residents and Clinical Fellows Grant (R25EB025787-01); the NIH Big Data to Knowledge (BD2K) Program of the National Cancer Institute (NCI) Early Stage Development of Technologies in Biomedical Computing, Informatics, and Big Data Science Award (1R01CA214825); NCI Early Phase Clinical Trials in Imaging and Image-Guided Interventions Program (1R01CA218148); an NIH/NCI Cancer Center Support Grant (CCSG) Pilot Research Program Award from the UT MD Anderson CCSG Radiation Oncology and Cancer Imaging Program (P30CA016672) and an NIH/NCI Head and Neck Specialized Programs of Research Excellence (SPORE) Developmental Research Program Award (P50 CA097007). Dr. Fuller has received direct industry grant support, honoraria, and travel funding from Elekta AB. S.J. Dr. Frank reports personal fees from Varian, grants and personal fees from C4 Imaging, grants from Eli Lilly, grants from Elekta, grants and personal fees from Hitachi, other from Breakthrough Chronic Care, personal fees from Boston Scientific, and personal fees from National Comprehensive Cancer Center (NCCN). Dr. Gunn reports philanthropic donation from the Family of Paul W. Beach.

